# A Spatial Analysis of Hypertension Prevalence in Rural and Urban Malawi

**DOI:** 10.1101/2021.04.23.21255979

**Authors:** James Burns, Mia Crampin, Alison Price, Chris Grundy, James Carpenter

**Author notes:** Corresponding author (. Department of Medical Statistics, The London School of Hygiene and Tropical Medicine, Keppel Street, London, WC1E 7HT).

## Abstract

**Background:** Non-communicable diseases (NCD) represent a large and rapidly growing disease burden in Malawi and the wider sub-Saharan Africa region. National and regional NCD prevalence estimates and mapping, establishment of associated risk factors, and trend monitoring are vital for sustaining hard-won gains in well-being and life expectancy from the successful management of infectious diseases.

**Methods:** Between 2012 and 2016, blood pressure was measured in a population-representative sample of 29,628 Malawian adults (18+ years) residing in two locations; the southern part of rural Karonga district and urban Area 25 in Lilongwe city. Each location was divided into approximately 200 zones with individuals assigned the zone in which their home was located. A conditional autoregressive (CAR) model was fitted to estimate zonal hypertension prevalence. Estimates were plotted on regional maps featuring key amenities, healthcare facilities, and transport links. Individual-level economic and lifestyle covariates were then incorporated to assess how much of the variation could be explained by those covariates.

**Results:** Variation in hypertension prevalence was observed in both the rural and urban location (*P* < 0.0005), with high-prevalence zones clustering in areas near to major transport links and/or concentrations of amenities. Covariates explained most of the variation in both sites (*P* > 0.14).

**Conclusions:** In rural and urban Malawi, hypertension prevalence is higher among those of relatively high wealth, those who are closer to local amenities, or a combination of these two factors. More detailed data are needed to determine if these associations are explained by wealth-consequent behaviours such as sedentary occupations and deleterious lifestyle choices.

## Introduction

Annually, 70% of global mortality is attributed to non-communicable diseases (NCD), amounting to over 41 million deaths of which 15 million are of individuals between 30 and 69 years. Over 85% of these premature deaths occur in low- and middle-income countries. Cardiovascular disease (CVD), cancers, chronic respiratory diseases and diabetes, are jointly responsible for 84% of worldwide NCD deaths (1).

The World Health Organisation’s 2030 Agenda for Sustainable Development recognises NCD as a major challenge for sustainable development in Africa, and predicts that over the next decade the continent will experience a 27% increase in NCD deaths, considerably higher than the predicted worldwide mean increase of 17%. Moreover, the WHO expects NCD deaths on the continent to exceed deaths due to communicable, maternal, perinatal and nutritional diseases combined by 2030 (2).

### NCD and Hypertension in Malawi

Malawi covers about 118,000 km^2^ (see Figure 1), has a population of roughly 18 million, and consistently ranks among the world’s poorest nations. NCD are estimated to account for 29% of deaths in the country, a figure that has been increasing year-on-year every year since 1990 when the figure was around 18% (3).

**Figure 1.**
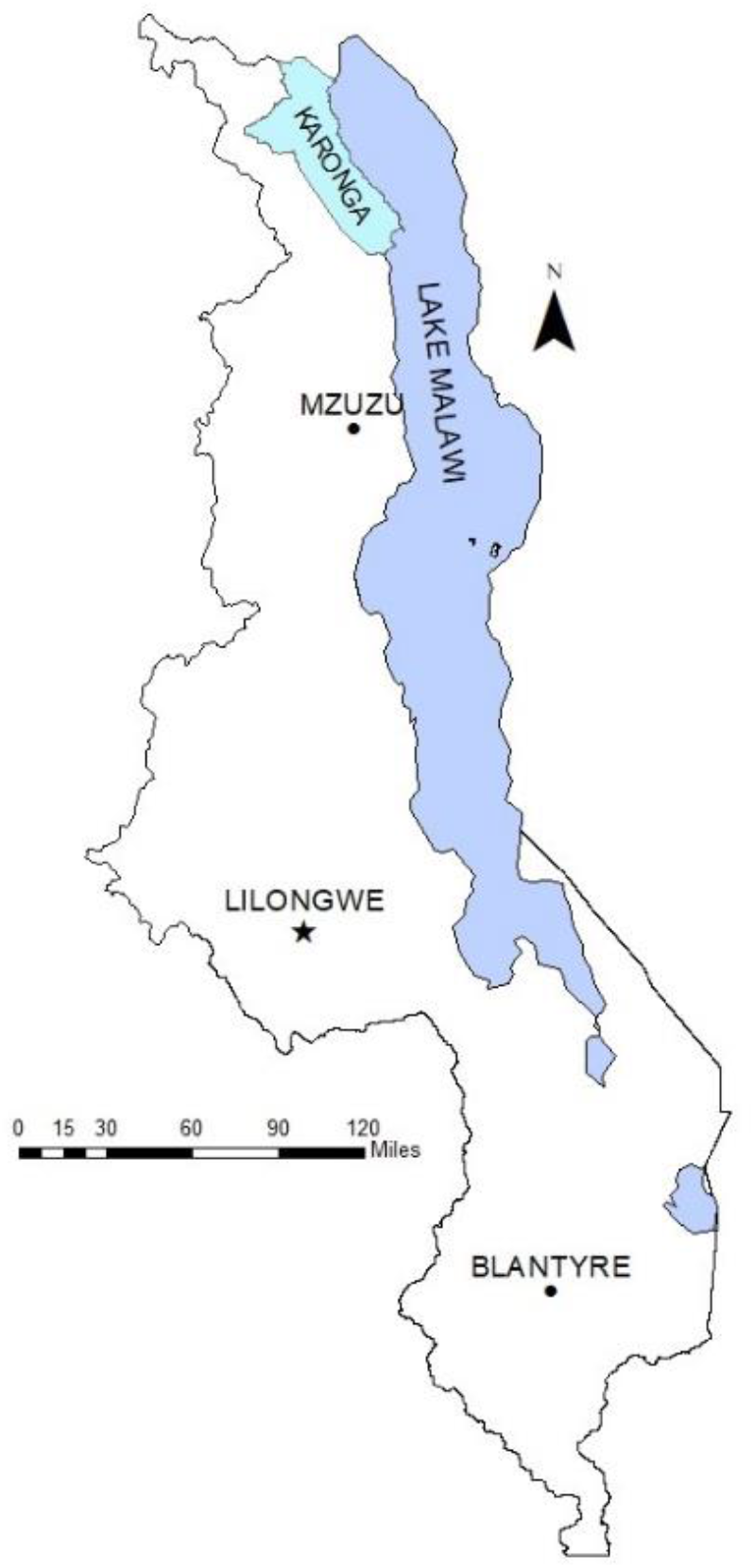
The nation of Malawi. The rural district of Karonga is shown in the far north, and the capital city of Lilongwe in the centre. These are the locations for the rural and urban study areas respectively.

In Malawi, hypertension is considered the strongest risk factor for CVD (4) which accounts for 12% of disability-adjusted life years lost to ill-health and early death (3). High prevalence of hypertension among urban over rural inhabitants has been shown in all age groups in Malawi (5), but the extent to which hypertension prevalence varies within urban and rural geographies, due to influences such as built environments and neighbourhood factors, is unknown. A spatial analysis of the prevalence of hypertension would inform future researchers and public health bodies as to the impact of these factors, so they can better develop and deploy public health measures designed to alleviate the disease.

### Links to Communicable Diseases

The interplay between communicable and non-communicable diseases has been demonstrated before, and the two are often linked. For example, HIV is increasingly recognised as a risk factor for CVD, with a meta-analysis of 20 studies reporting the relative risk of CVD, comparing HIV-infected patients to uninfected patients, to be 1.61. In the same study, patients taking anti-retroviral therapy to combat HIV had an even higher estimated relative risk, compared to those not, of 2.00 (6).

Conversely, NCD may increase the burden of communicable diseases. Bi-directional screening programmes that have been established in some low-middle income countries demonstrate that diabetes patients are over 3 times more likely to become infected with TB than those who do not suffer from the disease (7).

Given these links between communicable and non-communicable diseases, and the importance of spatial mapping to communicable disease epidemics (8), we hypothesise that the spatial structure of NCD epidemics can have an impact on the evolution of these epidemics and on the design of preventative and treatment interventions.

We investigate the extent to which spatial variation in the prevalence of hypertension exists in (a) a typical rural and (b) a typical urban location in Malawi, and whether any observed spatial variation can be explained by a range of socio-economic and lifestyle factors (e.g. smoking, alcohol intake, diet, physical activity, obesity, household wealth). We also investigate heterogeneity in these associations by location.

## Methods

NCD data used for these analyses, described in detail elsewhere (9), were collected by the Malawi Epidemiology and Intervention Research Unit (MEIRU) between 2013 and 2015 to determine prevalence of, and risk factors for, hypertension, diabetes and lipid abnormalities in rural and urban Malawi (9).

### Study Locations

The location of rural Karonga District, and urban Lilongwe, within Malawi are shown in Figure 1. In the south of Karonga district, the MEIRU operates the Karonga Health and Demographic Surveillance Site (HDSS) on the northern shore of Lake Malawi. The HDSS includes the village, port and trading centre of Chilumba, which hosts the HDSS headquarters.

The HDSS covers a population of approximately 44,000 (2018 estimate) (10) (11) most of whom depend on subsistence farming, fishing or trade for their livelihoods and who are spread over an area of roughly 135 km^2^. The HDSS is served by two hospitals: St. Anne’s Hospital on the northern perimeter and Chilumba Rural Hospital in Chilumba. There are also two health centres: Sangilo on the southern perimeter and Fulirwa on the western extremity of the east-west secondary road. There are several small clinics spread throughout the HDSS (12).

Area 25 is a rapidly growing urban area in the country’s capital and largest city, Lilongwe. The area is a mixed urban population of around 77,000 people (2018 estimate) (10) (11). Inhabitants range from civil servants, commercial and non-governmental organisation employees to students, living alongside large numbers of people in casual and informal employment such as petty trading or seasonal work at the local industrial estate, and those seeking work. Area 25 was selected to represent a typical c(11)ross-section of the inhabitants of urban Malawi. The site is roughly 15km from Kamuzu Central Hospital, the main tertiary hospital in Lilongwe (13).

### The Zones

Each area was divided into non-overlapping zones that jointly covered it. There were 209 zones in the Karonga site, containing an average of 36 households each (range 11 to 101), and 200 zones in Area 25, containing on average 42 households each (range 28 to 88). Mean zonal study participants were 69 in Karonga (range 13 to 180), and 76 in Area 25, Lilongwe (range 43 to 170).

### Data Collection

Participants were visited in their homes and written informed consent was obtained for a questionnaire, anthropometry, and blood samples. All adults (18+) were eligible, and households were visited at least three times (including weekends) to recruit eligible individuals missed at earlier visits.

For each area, the geoposition of participants’ dwellings were captured along with co-ordinates of major and minor roads, tracks, rivers and key institutions and amenities (such as health facilities and schools).

### Blood Pressure Measurements

For all individuals, the blood pressure measurements were conducted as follows. After thirty minutes of rest, three seated blood pressure measurements, with five minutes rest between each, were collected on the right arm when possible (otherwise collected on the left arm in those with conditions that precluded used of the right arm) using portable sphygmomanometers (OMRON-Healthcare-Co HEM-7211-E-Model-M6; Kyoto, Japan). The mean of the last two blood pressure readings was used (13). Individuals were classed as hypertensive if they had a systolic blood pressure measurement in excess of 160 mmHg, or a diastolic blood pressure measurement in excess of 100 mmHg.

**Table 1.** Tabulations of the outcome of hypertension, along with other covariates, by setting.

| Variable: | Karonga (Rural)<br>count (%) | Lilongwe (Urban)<br>count (%) |
| --- | --- | --- |
| **Hypertensive:** |  |  |
| Yes | 1,695 (12.2%) | 2,133 (12.8%) |
| No | 12,197 (87.8%) | 14,529 (87.2%) |
| **Sex:** |  |  |
| Male | 5,864 (42.2%) | 5,804 (34.8%) |
| Female | 8,039 (57.8%) | 10,867 (65.2%) |
| **Age:** |  |  |
| mean (SD, range) | 38.0 (16.4, 17 to 105) | 32.4 (12.9, 17 to 103) |
| **Possessions Score:** |  |  |
| 1 (least wealthy) | 5,098 (36.7%) | 3,041 (18.2%) |
| 2 | 6,495 (46.7%) | 5,829 (35.0%) |
| 3 (wealthiest) | 2,310 (16.6%) | 7,802 (46.8%) |
| **Electricity:** |  |  |
| Yes | 1,107 (8.0%) | 9,449 (56.7%) |
| No | 12,796 (92.0%) | 7,223 (43.3%) |
| **Water Source:** |  |  |
| Tap to house | 2,782 (20.0%) | 10,432 (62.6%) |
| Other | 11,121 (80%) | 6,240 (37.4%) |
| **Alcohol Intake:** |  |  |
| Never | 11,154 (80.2%) | 13,837 (83%) |
| Rarely | 1,542 (11.1%) | 1,822 (10.9%) |
| Often | 1,207 (8.7%) | 1,013 (6.1%) |
| **Overweight:** |  |  |
| No | 10,794 (80.8%) | 10,445 (64.9%) |
| Yes | 2,562 (19.2%) | 5,649 (35.1%) |

### Measuring Modifiable Exposures

Covariates in this analysis included age, sex and modifiable exposures wealth, overweight and alcohol intake. These were chosen as preliminary analysis demonstrated associations between them and hypertension in the data (see Model Fitting below).

Anthropometry measurements including weight and height were measured twice by two study staff independently, following removal of shoes and outer clothing, using calibrated Seca scales, stadiometers, and flexible tape measures. The means of the two measurements were used in these analyses to calculate body mass index (BMI), with overweight/obese defined as a BMI ≥ 25kg/m^2^.

Information on age, sex, wealth and alcohol intake were collected using an interviewer-led questionnaire. Alcohol intake was self-reported and grouped as never, rarely (on at most 4 days a week) and often (on more than 4 days a week). Water source (house tap/no house tap) and electricity access (yes/no) were used as proxies for wealth, but as these are more a function of location rather than wealth in the rural area, the primary means of estimating household wealth was mean household assets score.

For this, a list of assets owned by each household was compiled, and the local price of each estimated by staff with knowledge of the setting. Each household was then assigned an assets score based on these values. Scores from both sites were merged and sorted into deciles which were then grouped into 3 categories, deciles 1-3, 4-7 and 8-10 and named 1, 2 and 3 respectively (3 being the highest mean possession score).

### Model Fitting

Each zone was given a ‘neighbourhood’ defined as the set of zones with which it shared a border, excluding the zone itself.

#### Unadjusted Analysis

First, a standard mixed-effects model (parameters fitted using ML) was run, with no covariates, and compared to a corresponding fixed-effects-only model by means of a likelihood ratio test. The *P*-value for the significance of the variance of the random effect estimate, quantified the evidence against the hypothesis that there is no zonal variation in hypertension.

Next, a cross-classification, conditional auto-regressive (CAR) model was used to model hypertension prevalence in each zone. This is a mixed-effects model, where each zone is assigned a random effect from its own unique distribution, and the parameters of this distribution are derived from the zone’s neighbourhood. This model was chosen as it was felt it would account for anticipated correlation of hypertension prevalence between nearby zones; it aims to give a clearer picture of the underlying hypertension prevalence pattern by borrowing data from each zone’s neighbours and incorporating this into each zone’s estimate. In particular, the mean of each zone’s random effect distribution was the mean prevalence of its neighbourhood, and its variance was the global random effect variance divided by the number of zones in the neighbourhood:

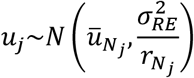

Where *u*_*j*_is the random effect for zone *j*, 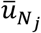 is the mean prevalence of zone *j*’s neighbourhood, 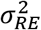 is the global random effect variance, and 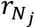 is the number of zones in *j*’s neighbourhood.

This led to distributions centred on the neighbourhood mean prevalence, which were more precise if the zone had many neighbours, and less precise if it had few. The *P*-value for the global random effect variance, 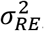, was used to determine the statistical significance of the variation of hypertension prevalence across zones.

This model has the effect of ‘smoothing’ the prevalence estimates, aiming to mitigate the fact that we have imposed arbitrary boundaries on an underlying process that we believe to be evolving continuously over space.

The CAR model was fitted using Markov Chain Monte Carlo (MCMC) methods, where Gibbs sampling was used to sample from parameter posterior distributions. Diffuse priors were used for variance parameters. In particular: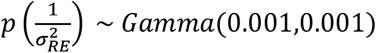, which excludes 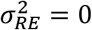 as a value. The iterative generalised least squares (IGLS) algorithm was used to inform starting values for parameter chains – two chains were run with starting values two standard errors above, and two below, the estimates arrived at by IGLS. Estimates were based on 20,000 samples with a 500-sample burn-in.

#### Adjusted Analysis

The adjusted analysis investigated how much of the spatial variation in hypertension prevalence could be explained by socioeconomic and lifestyle covariates. The covariates considered were age, sex, wealth, overweight and alcohol intake. These were chosen based on standard logistic regression analysis which showed them to have an association with the hypertension status. Information on salt intake (a key risk factor for hypertension) was collected but left out of the analysis; a 2018 paper which used the same data as this analysis found that there was no correlation between the household salt intake as measured by the method used in collecting this data and an individual measurement derived from collected spot urine samples (14).

Covariates were added to the unadjusted model just outlined, and from this we derived an estimate of each zone’s marginal hypertension prevalence using only the fixed effects in this model. The difference, *d*_*j*_, between this estimate and the observed hypertension prevalence for each zone was plotted, using a three-colour gradient, and the mappings used to determine how much the covariates explained the observed variation. For comparison, the same was done for a model with no covariates. (Note that this simply reduced to plotting the difference between the zone’s observed hypertension prevalence and the global mean). If the covariates explained much of the variation, we might expect previously identified spatial patterns to disappear when this new measure is plotted. If not, we would expect those patterns to remain.

*χ*^2^-like statistics were calculated for each zone, for each model, and their sum compared to an appropriate *χ*^2^ distribution:

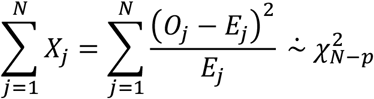

Where *O*_*j*_and *E*_*j*_are the observed and expected (by the relevant model) hypertensive counts for zone *j. N* is the number of zones and *p* is the number of parameters in the model. Models were fitted using the same MCMC methods as for the unadjusted analysis.

## Results

Data on hypertension status and all considered covariates were available for 13,892 individuals in rural Karonga and 15,736 in urban Lilongwe. All were used in the models.

### Unadjusted Models

The modelling suggested very strong evidence for spatial variation in hypertension prevalence in rural Karonga. The variance of the random effect, 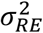, was estimated to be 0.30 (*P* = 0.0001) with a 95% credible interval of 0.16 to 0.50.

Figure 2 shows the mapping from the unadjusted CAR model for rural Karonga. On the same figure, the mean household possession score is also plotted for each zone, and the geographical association between higher mean possession score and high estimated hypertension prevalence is visible.

**Figure 2.**
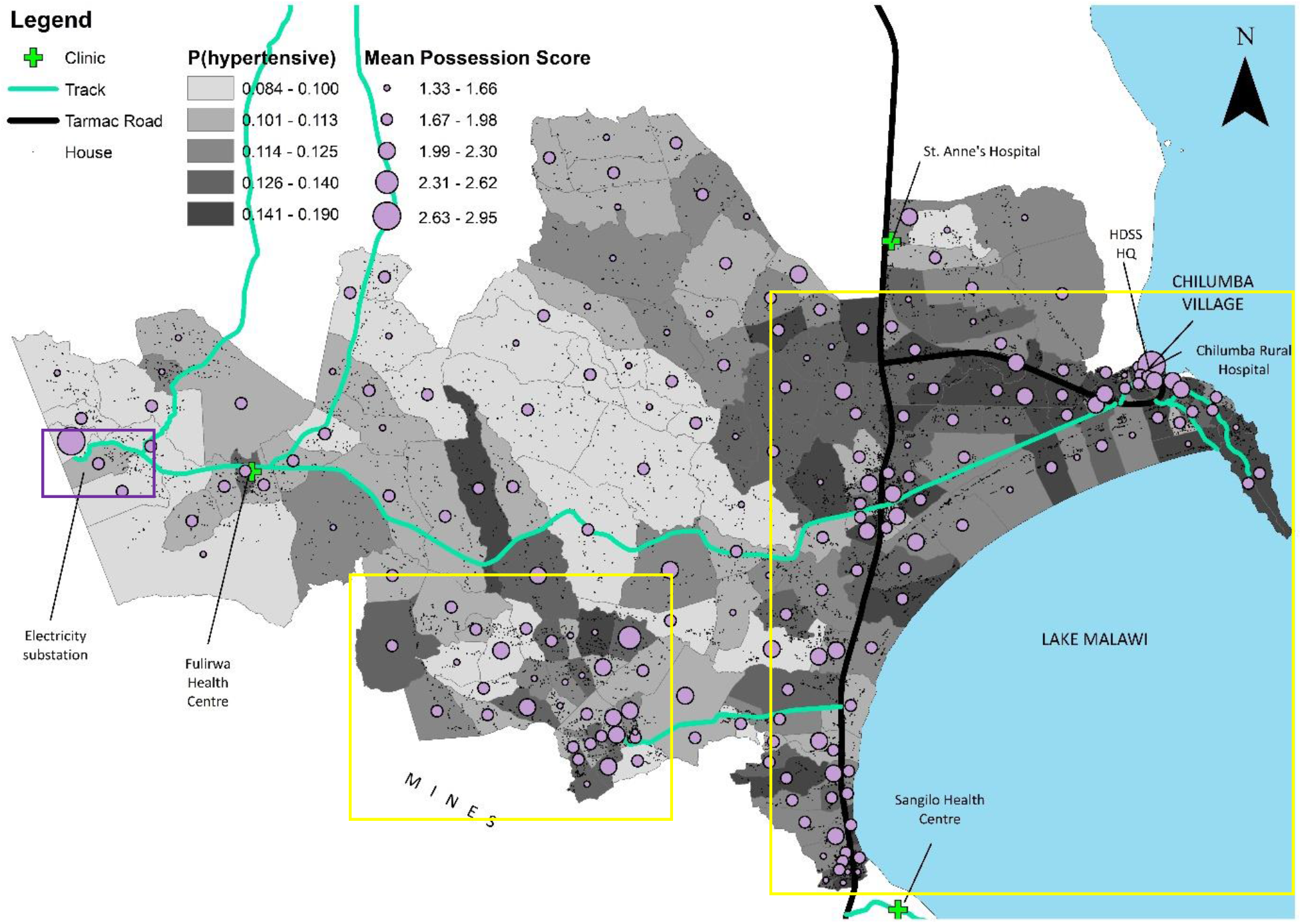
Mapping for Karonga. Zone shading indicates predicted marginal probability of an individual being hypertensive, with darker shade indicating higher probability. The circles represent zonal mean household possessions score, with larger circles indicating higher mean possession score. HDSS stands for ‘Health and Demographic Surveillance Site’.

We see several areas where high predicted probability of hypertension appears to be concentrated. The high-risk coastal triangle formed by the shore of Lake Malawi and the east-west and north-south tarmac roads (large yellow box) contrasts strikingly with the rest of the region. The southern boxed area, close to mining industry off-map to the south, has arguably emerged as a second area of high hypertension risk relative to outlying areas. The zone containing a remote electricity substation (purple box) is in a higher hypertension probability category than all its neighbours.

The seemingly anomalous central zone with highest risk classification just above the smaller yellow box (zone 111) contains 84 study participants, more than 77% of zones, 19 of whom are hypertensive. This proportion (0.23) is the tenth highest of any zone and is considerably higher than the site-wide average of 0.12. It is unclear what is driving this.

In urban Area 25, Lilongwe, there was also very strong evidence for spatial variation in hypertension prevalence. The variance of the random effect, 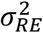, was estimated to be 0.028 (*P* < 0.0001) with a 95% credible interval of 0.013 to 0.049.

Figure 3 shows the mapping for Area 25, Lilongwe. Firstly, there is no obvious coincidence between high predicted hypertension prevalence and higher income. Secondly, we see that almost all zones with the highest predicted probability of hypertension are in the south-central and south-east regions of the area, along with most of the major roads and facilities, and in closest proximity to the centre of Lilongwe (located south of Area 25). West of the north-south road, all zones except three are in the lowest two classes of random effect estimate, and everywhere else we see mostly mid-range measures.

**Figure 3.**
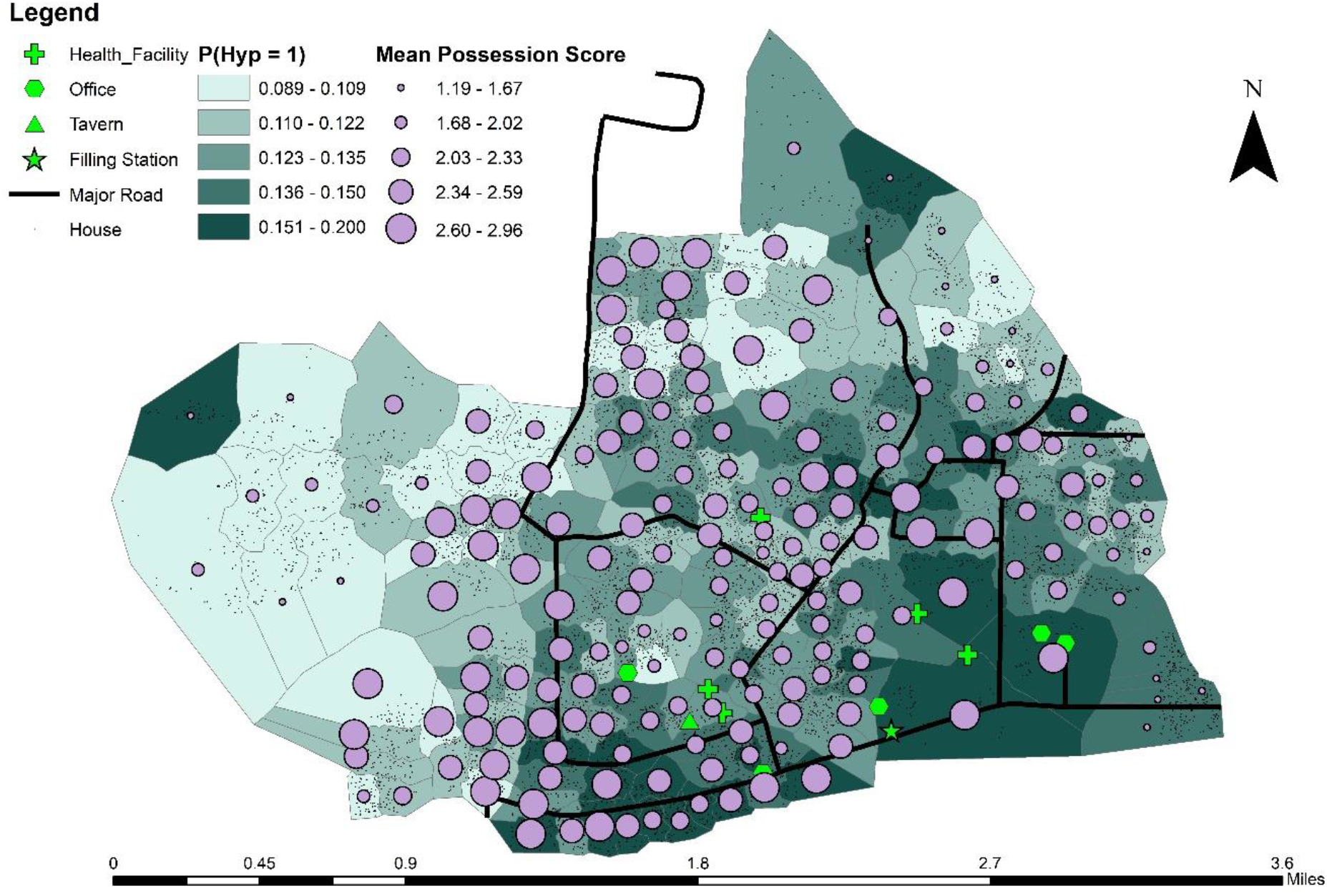
Mapping for Area 25, Lilongwe. Zone shading indicates predicted marginal probability of an individual being hypertensive, with darker shade indicating higher probability. Circles represent mean zone possession score, with larger circles indicating higher mean possession score.

The outlying highest-risk zone on the north-west border (zone 72) contains fewer study participants than average (43 compared to a mean of 76) and has an observed hypertensive proportion of 0.14, which is close to the site-wide average of 0.13. As such, and considering it is surrounded by lowest-risk zones, it is unclear why the modelling has classified this as a highest-risk zone.

The other outlying highest-risk zone on the north-east border (zone 2) has a greater-than-average population of study participants at 81 (average is 76), and has the highest observed proportion of hypertensives of all zones at 0.26. It is unclear what is causing this high hypertension prevalence.

### Adjusted Models

In Karonga, the approximate *χ*^2^ statistic for the unadjusted model was 259.4 (*P* < 0.009), compared to 221.7 (*P* = 0.14) for the adjusted model, suggesting that the covariates have explained much of the variation in hypertension prevalence across the site. Kernel density plots also indicate the differences, *d*_*j*_, are more concentrated around zero for the adjusted model (see Figure 6 in the appendix).

Figure 4 (top) shows, in Karonga, the differences, *d*_*j*_, between the modelled unadjusted hypertension prevalence and the observed prevalence for each zone. Beneath this is the equivalent figure for the covariate-adjusted model. On each, the *χ*^2^ statistic contribution for each zone is also plotted. We can see that the covariates have explained some of the hypertension variation in the coastal triangle (formed by the lakeshore and the two tarmac roads, yellow box) where there was very high hypertension prevalence. However, it has not explained as much in the south-central region (red box) adjacent to the mines; here the plots of the *d*_*j*_ are more or less the same for both models, suggesting that there might be factors which are not captured by the covariates in our adjusted model.

**Figure 4.**
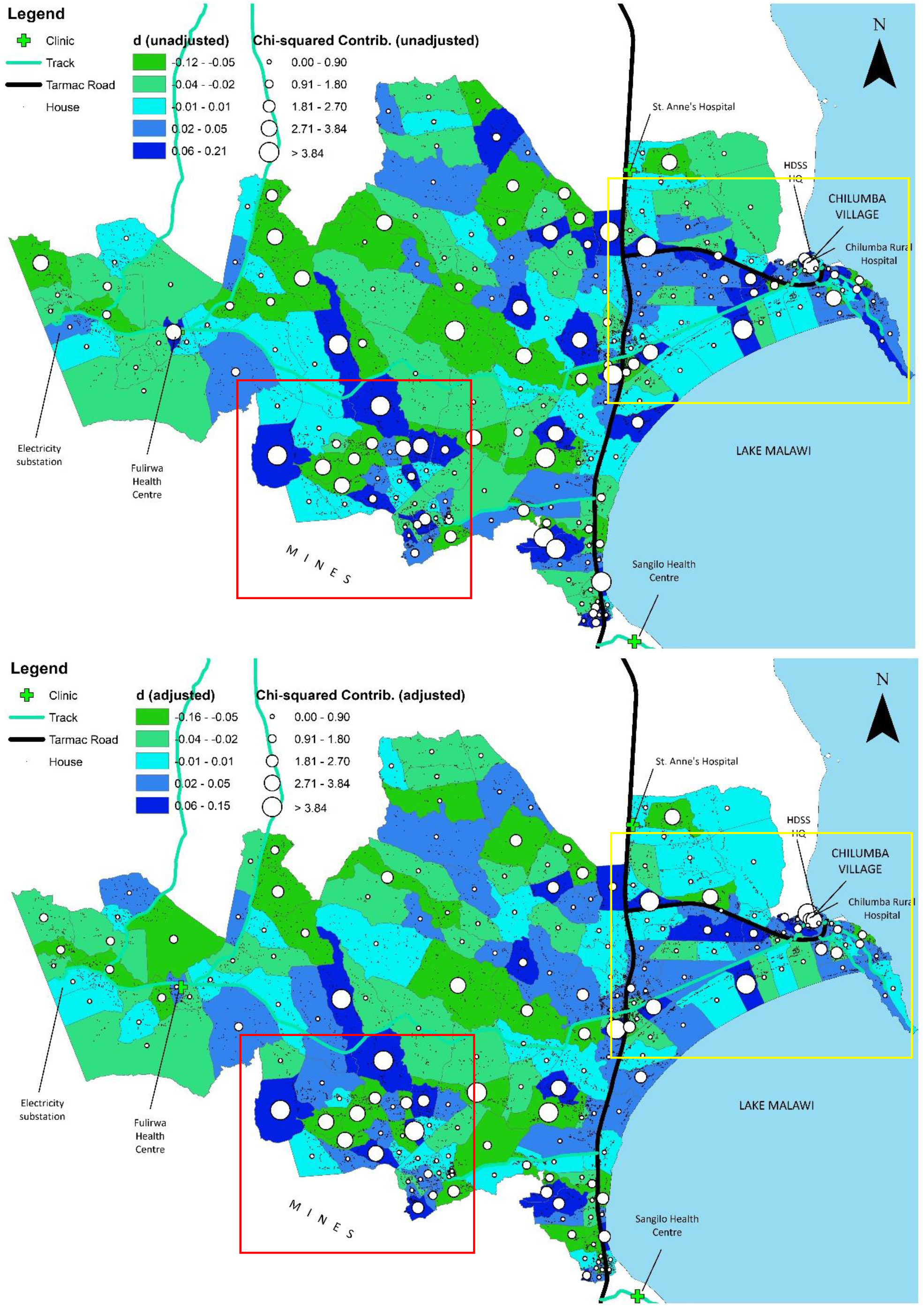
Mappings of the differences, *d*_*j*_, for the unadjusted model (top) and covariate-adjusted model (bottom) in Karonga. *d*_*j*_ is the difference between the observed hypertension prevalence and our covariate-based estimate for zone j. The circles represent the corresponding contribution to the approximate *χ*^2^ statistic for each zone.

Concerning the urban site in Area 25, Lilongwe, the approximate *χ*^2^ statistic for the unadjusted model was 272 (*P* < 0.002), compared to 173.3 for the adjusted model (*P* = 0.91). Kernel density plots also indicate the differences are more concentrated around zero for the adjusted model (see Figure 6 in the appendix). This again suggests that the covariates have explain much of the variation in hypertension prevalence across the site, leaving only variation compatible with chance.

Figure 5 shows mappings of the differences, *d*_*j*_, between the modelled unadjusted hypertension prevalence and the observed prevalence for each zone. Beneath it is the same but for the covariate-adjusted model prevalence estimates. On each, the *χ*^2^ statistic contribution for each zone is also plotted. From inspecting the plot, it is not clear where precisely the reduction in variation has occurred. Some of the variation in the north boxed area appears to have been explained, however little of that in the south-central region (smaller box) and the south-east corner appears to have been explained.

**Figure 5.**
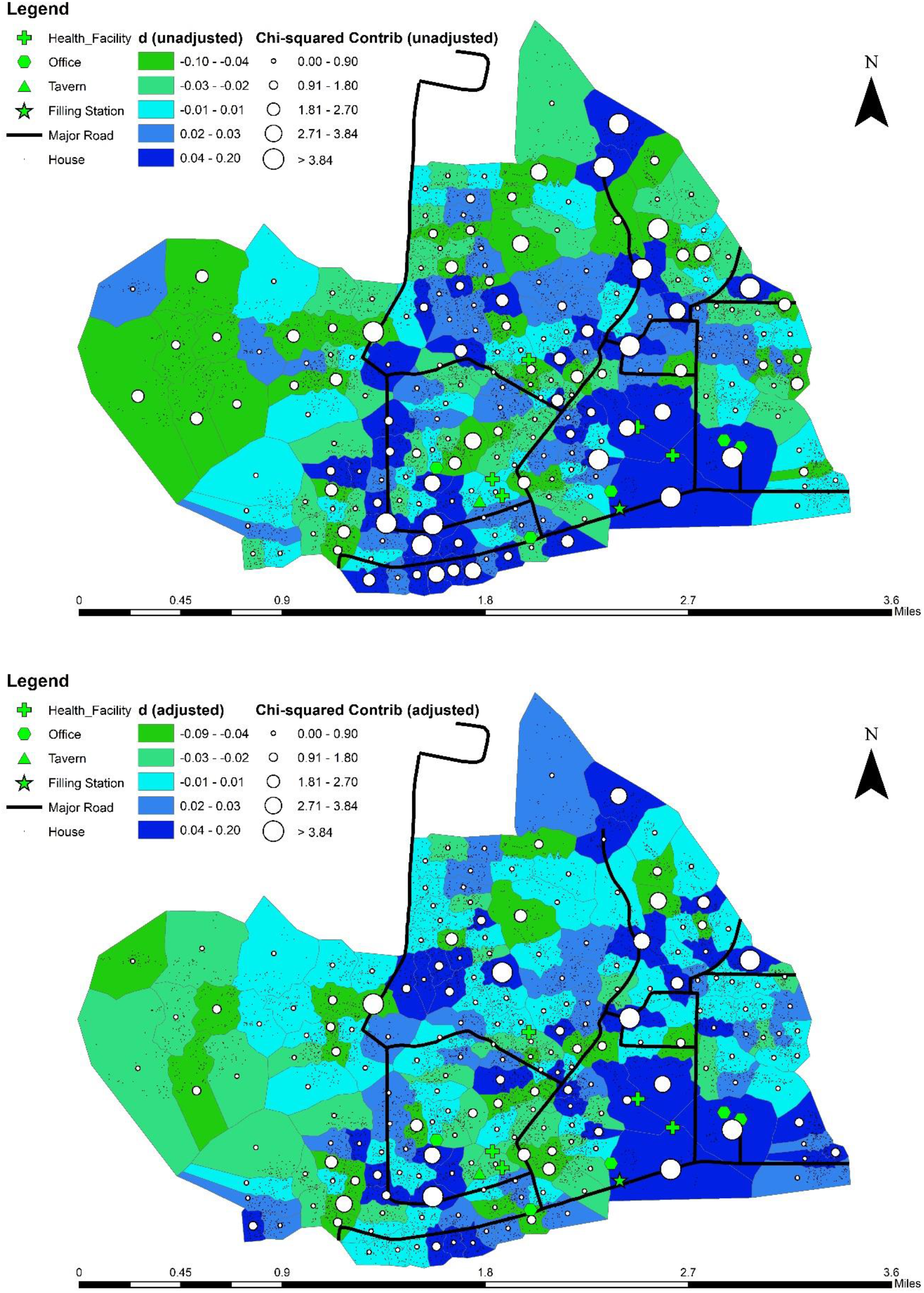
Mappings of the differences, *d*_*j*_, for the unadjusted model (top) and covariate-adjusted model (bottom) in Lilongwe. *d*_*j*_ is the difference between the observed hypertension prevalence and our covariate-based estimate for zone j. The circles represent the corresponding contribution to the approximate *χ*^2^ statistic for each zone.

## Discussion

Spatial clustering of hypertension was seen in both rural and urban settings in low-income Malawi and, similarly to neighbouring, middle-income South Africa, this was largely explained by clustering of individual risk factors (13).

In both locations, higher prevalence of hypertension was associated with more ‘developed’ areas that had (a) access to key regional transport links, (b) a high concentration of facilities relative to the rest of the region, (c) proximity to urban/industrial centres, or (d) some combination of these. It is possible that these factors result in greater exposure to goods and conveniences (e.g. processed and prepared foods) or allow for a more sedentary lifestyle conducive to the development of hypertension. It could also be that people who are more likely to make these lifestyle choices choose to live in proximity to these areas.

In particular, in Karonga, the east-west tarmac road extends to the jetty and lakeside (top yellow box in Figure 2) with an untarred road branching northward to the HDSS headquarters. Residents living on this ‘jetty’ road are more likely to have formal employment, and easier access to other parts of the region and the country, relative to more remote residents. The concentration of high hypertension prevalence also continues (but is a little less pronounced) along the north-south tarmac road, where lifestyles and transport links are more likely to be similar compared to neighbourhoods far from these two roads.

The observed higher hypertension prevalence in the area close to the mining industry (bottom yellow box on Figure 2) was not explained by it being a relatively high-income area; only 2.4% of participants in this area were employed compared to 7% in the coastal triangle and 3% elsewhere. However, participants in this zone were less likely than participants in outlying zones to be subsistence farmers or fishermen (69% vs. 78%) and more likely to be self-employed (12% vs. 6%). Participants here were more than three times more likely to have electricity in their home than in outlying areas (7% vs. 2%). However, because electricity availability is so strongly determined by precise location in Karonga (most importantly: proximity to the main roads) electricity access cannot be considered a direct signifier of wealth. Yet it is also likely that electricity availability is a desirable amenity in Karonga, and consequently possible that only wealthier households can afford to live in areas that have a higher proportion of electrification than outlying areas.

In both locations covariates (specifically age, sex, wealth, overweight and alcohol intake) explained much of the spatial variation in hypertension, leaving only random variation compatible with chance (*P* ≥ 0.14). Once controlling for these covariates, reduction in high-hypertension cluster patterns was visible on the plots in Karonga but not in Lilongwe. We posit that this might be because Area 25 is more homogenous in terms of the covariates that Karonga, and thus the variation reduction has occurred site-wide in a uniform way.

A clear association between higher household development and higher hypertension prevalence was observed in rural Karonga but not urban Lilongwe. This difference may be because a greater proportion of urban Area 25 residents live in more-developed households with a lower spread of development measures compared to rural Karonga. Notably, in Area 25, 55% of individuals had electricity access compared to only 7% in Karonga. Furthermore, the mean possessions score was 2.3 (SD = 0.36) in Area 25, compared to 1.7 (SD = 0.22) in Karonga. In Area 25, 80% of zones had a mean possession score of 2 or higher compared to just 15% in Karonga. We might posit that in rural Karonga, those with higher household development have some disposable income leading to deleterious lifestyle choices, whereas in Area 25 it is proximity to facilities, and subsequent ease of partaking in deleterious lifestyle choices, that is driving the pattern as opposed to local relative wealth and white-collar employment.

The main limitation of the study is that, while it did establish the extent to which lifestyle covariates explained spatial variation in hypertension prevalence, it could not establish *which* lifestyle factors were associated with *which* regions of high hypertension prevalence (including, notably, sodium intake) and further studies should seek to address it.

‘Hotspots’ of high hypertension in highly populated and amenity-rich areas were identified in both areas which could potentially provide a focus for campaigns to reduce risk and improve ascertainment.

## Ethics Approval

Ethical approval for the study was granted by the Malawi National Health Sciences Research Committee (NHRSC) protocol number #1072, and LSHTM Ethics Committee protocol number #6303.

## Data Availability

Data is owned and controlled by the Malawi Epidemiology and Intervention Research Unit.

