## Supplemental_figure for "A Spatial Analysis of Hypertension Prevalence in Rural and Urban Malawi"

### Supplementary materials

Figure 6. Kernel density plots showing the distribution of differences, d_j_, resulting from the unadjusted and adjusted analyses.

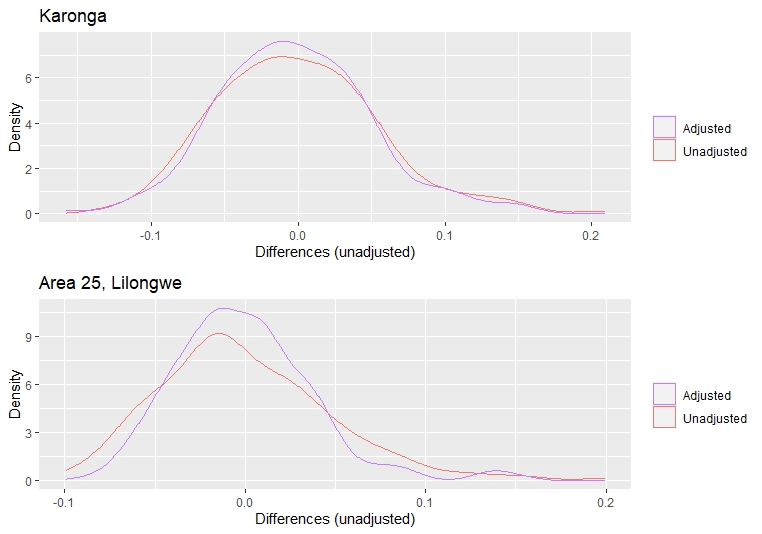
